# CladePredictor–MPXV: An alignment-free Artificial Intelligence-based classifier of complete and partial mpox virus genomes

**DOI:** 10.64898/2026.04.27.26351821

**Authors:** Gustavo Sganzerla Martinez, Ali Toloue Ostadgavahi, Mansi Dutt, Finlay Maguire, Lourdes Peña-Castillo, David J Kelvin, Anuj Kumar

**Author notes:** Correspondence: David J Kelvin.

## Abstract

Poxviruses constitute a threat to human health. Since 2022, two public health emergencies of international concern due to global spread of mpox viruses (MPXVs) were declared. The emergence of the novel MPXV subclade Ib has placed the global health community on alert as sustained human-to-human and travel-related transmission is prevalent in Africa and 30 non-African countries. Metagenomic and outbreak surveillance data often generates complete as well as partial assemblies of genomes which then require efficient taxonomic classification. Traditional viral genome classifiers rely on poorly scalable alignment methods creating computational bottlenecks in taxonomic classifications. Here, we present CladePredictor– MPXV: an alignment-free AI-based classifier of complete and partial MPXV genomes. Our classification framework consists of an ensemble of XGBoost and CNNs to classify between subclades Ia, Ib and IIb. CladePredictor–MPXV was trained with 3,866 MPXV genomes. XGBoost models were trained with 3-mers which are representative of the global feature space of complete MPXV genomes. CNNs were trained with short-range, position-independent sequence patterns to assign clades to partial genomes with a minimum size of 1000 nucleotides. Our XGBoost instance attained a weighted average accuracy of 90.2% while our CNN instance attained a weighted average accuracy of 95% in classifying clade (I vs II) and subclade (Ia vs Ib) from complete (>= 188,000 nucleotides) and partial MPXV genomes on a phylogenetically distinct validation set. CladePredictor-MPXV is freely available at https://clade-predictor.microbiologyandimmunology.dal.ca and provides a fast and efficient framework for the assignment of clades to MPXV subclade Ia, Ib, and IIb complete and partial genomes.

## INTRODUCTION

Mpox (formerly monkeypox) is a viral zoonotic disease caused by the mpox virus (MPXV) that is endemic to West and Central Africa. MPXV is a species of the genus *Orthopoxvirus* and is divided into two clades: clade I (also known as Central African Clade) and clade II (also known as West African Clade) [1]. In 2022, the MPXV subclade IIb emerged in countries with no prior history of mpox outbreaks. Sustained human-to-human transmission of this subclade led to the World Health Organization (WHO) declaring a Public Health Emergency of International Concern (PHEIC) [2]. In September 2023, the novel MPXV subclade Ib was identified in eastern DRC [4,5,6]. Spread of MPXV subclade Ib to neighboring countries, and incidences of clade I viruses emerging outside the African continent led the WHO to declare its second mpox-related PHEIC in less than three years. Kinshasa, the capital of DRC, is one of the most populated metropolitan areas in the African continent (∼17 million people) and is simultaneously experiencing mpox epidemics caused by subclade Ia and Ib viruses [7]. Both clades demonstrated capacity for sustained human-to-human transmission [6,20]. In addition, despite limited epidemiological data to ascertain clinical differences between the diseases caused by the two clades in humans [21], evidence in animal models suggests higher pathogenicity associated with clade I viruses [22]. The eastern region of the Democratic Republic of the Congo (DRC) has been experiencing armed conflict for decades, which has caused over 1 million Congolese seeking refugee abroad and 21 million people in the country in urgent need of food, medical, and other aid [3]. The forced displacement of people due to war can lead to mass migrations of already stressed populations into new ecozones, increasing contact with local wildlife that may harbor unfamiliar pathogens. Such interactions elevate the risk of zoonotic spillover events [19].

The ongoing spread of mpox highlights the importance of understanding genetic differences between its subclades, which can vary in terms of transmission dynamics, geographic distribution, and clinical outcomes. During outbreak situations, biomedical research centers generate large volumes of sequencing data. To translate this data into actionable insights, there is the need for tools capable of classifying viral subtypes both quickly and accurately. Traditional MPXV subtyping tools, such as NextClade [8] and Genome Detective [9], rely on sequence alignment algorithms. While these methods are well-established for comparing sequences of varying lengths, they are computationally expensive due to their pairwise or multiple sequence alignment steps, which often involve dynamic programming algorithms with time and space complexities of at least O(n^2^) or O (nm. As a result, their scalability is limited when processing large datasets of complete viral genomes or when rapid classification is required in outbreak settings. In this context, alignment-free methods are advantageous. Some examples of alignment-free tools are Kraken2 [10], a taxonomic classification system that uses k-mer matches to classify genomes, and Craft [11], which consists of a Machine Learning (ML) framework for classifying dengue subtypes. In fact, ML-based algorithms are promising methods to identify patterns in non-linearly related data [12,13,14].

Here, we present CladePredictor-MPXV: an alignment-free AI-based classifier of complete and partial MPXV genomes in the format of a user-friendly web server (https://clade-predictor.microbiologyandimmunology.dal.ca) that enables complete or partial MPXV genomes to be rapidly classified into their clades (I and II) and subclades (Ia and Ib).

## MATERIALS AND METHODS

### 2.1 Mpox virus genomes

We obtained a total of 3,866 MPXV complete genome sequences from the Global Initiative on Sharing All Influenza Data (GISAID) [15] deposited from 2001-08-08 to 2025-08-08. The data is divided into 646 MPXV subclade Ia, 593 subclade Ib, and 2,627 subclade IIb genomes. To account for phylogenetic structure in the dataset and avoid overestimating model performance due to closely related sequences being present across data splits, we applied a phylogenetically stratified sampling strategy. All genome sequences were processed using mash, a tool for genome and metagenome distances estimation based on MinHash sketches. A sketch of each sequence was created using the default parameters (k=21, sketch size = 1000) and pairwise distances were calculated using mash dist. The resulting mash distance matrix was then parsed into a symmetric square form and subjected to hierarchical clustering using average linkage. A distance cutoff of 0.05 was applied to define clusters of closely related sequences. These clusters were interpreted as approximate phylogenetic groups used to guide the data portioning into training, testing, and validation sets, ensuring sequences from the same phylogenetic cluster were not split across different subsets. Following Mash distance clustering, we identified a dominant cluster containing ∼98% of the sequences (555 subclade Ia, 584 subclade Ib, 2,627 subclade IIb). To simulate emerging clade detection, we used random 70% of the major cluster for training, random 15% for testing, and random 15% for validation together with all sequences from minor clusters. The train data is composed by 392, 392, and 1852, the test data is composed by 80, 96, and 388, while the validation data is composed by 173, 105, and 387 MPXV subclade Ia, Ib, and IIb genomes, respectively. The subclade composition of each cluster is available in **Supplementary Materials S1**.

### 2.2 Feature engineering

We used different approaches to encode complete (>= 188,000 bps) and partial MPXV genomes. First, each complete genome was encoded into a matrix of 3-mers frequency where we aimed to capture global features of complete genomes. The 3-mer matrix accounts for every possible combination of triplets of non-ambiguous nucleotides (ATCG), totaling 64 3-mers. Second, to classify partial MPXV genomes, each genome was split into *n* windows of 1000 nucleotides each in a 500-nucleotide step of overlap. Each 1000-nucleotide window was converted into a one hot encoding vector (A:1000; T: 0100; C: 0010; G:0001). The data matrices were built using in-house Python (version 3.12) scripts with the Python modules pandas (version 2.2.3), Bio (version 1.85), collections (version 3.12), and itertools (version 3.12).

### 2.3 2-D visualization of trinucleotide frequency of MPXV genomes

We visualized in matplotlib (version 3.10) and seaborn (version 0.13.2) each of the genomes separated into subclades Ia, Ib, and IIb containing 64 dimensions (3-mer frequency) in a 2-D plane using Uniform Manifold Approximation and Projection (UMAP, Python umap version 0.5.8) iterating over the UMAP parameters number of neighbors (1%, 5%, and 10% of the observations in datasets), minimum distance (0.1, 0.3, 0.5, 0.7, and 1), and distance metric (Euclidean, Manhattan, and Cosine). We calculated the Silhouette score of each iteration to quantify how well-separated the classes are in the UMAP plots (the Silhouette score closer to +1 was selected as it represents the best-defined clusters).

### 2.4 XGBoost classification of complete MPXV genomes

Complete subclade Ia (646), Ib (318), and IIb (2,627) MPXV genomes obtained from GISAID were encoded into trinucleotide (3-mer) frequency and used to create a dataset for classification. We trained two instances of an XGBoost classifier. We used the sklearn (version 1.6.1) python module implementation of each XGBoost classifier. Two classification tasks were considered, i.e., clade (clade I vs. clade II) and subclade (subclade Ia vs. subclade Ib). Both XGBoost models were instantiated with 200 boosting rounds with a learning rate of 0.1. The maximum depth of each decision tree was set to 6. The models employ subsampling (subsample = 0.8) and column sampling (colsample_bytree=0.8) to introduce randomness. The objective function was “binary:logistic” optimized using the log loss metric. Each XGBoost model was assessed as per its accuracy, precision, recall, and F1-score over the train, test, and validation datasets.

### 2.5 XGBoost classification of partial MPXV genomes

The XGBoost models for clade and subclade trained with complete genomes were stressed in classifying partial MPXV genomes. To assess the classification performance of genome fragments of varying lengths, partial genome sequences were systematically generated from the full-length sequences in the validation dataset. For each genome sequence, random sub-sequences were extracted at lengths varying from a maximum length *L*_*max*_ = 196,000 base pairs down to a minimum length *L*_*min*_ = 175,000 base pairs, decrementing in steps of ΔL = 1,000 base pairs.

For each partial genome length *L* such that:

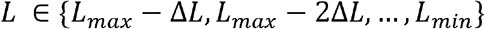

A sub-sequence of length *L* was randomly sampled from the full genome sequence of length *s* (where *s* ≥ *L*_*max*_) at random start position *p*, uniformly drawn from

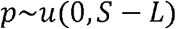

The extracted partial genome *G*_*L*_is thus

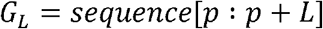

This process was repeated for all genome sequences in the validation dataset to generate sets of partial genomes at each length *L*. Each *G*_*L*_ was then encoded using 3-mers frequency vectors and used as input for the XGBoost classification models of clade and subclade genomes, enabling performance evaluation across a range of partial genome sizes.

### 2.6 Generation of partial MPXV genomes

To prepare the MPXV sequences for CNN classification, each genome sequences was partitioned into overlapping windows of fixed length *W*= 1000 base pairs. The windows were generated by sliding a fixed-size window across all the complete genomes with a step size *S* = 500 base pairs, producing a set of subsequences {*G*_*i*_} defined as:

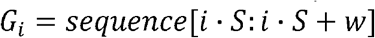

Where *i* = 0, 1, 2, …, *N* and 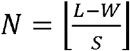, with *L* being the length of the full genome sequence. Each window *G*_*i*_ was encoded using one-hot encoding to transform nucleotide characters into a numerical vector format suitable for CNN input. The nucleotide encoding scheme is defined by a dictionary mapping nucleotides to orthogonal vectors:

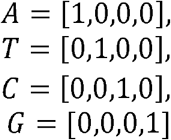

Each window *G*_*i*_, consisting of a nucleotide sequence of length *W*, is transformed into a matrix X_*i*_ ∈ ℝ^*Wx*4^, where each row is the one-hot encoded nucleotide vector. This process was applied to all sequences across train, test, and validation datasets. Class labels corresponding to each sequence’s subclade was assigned to all its windows, forming labeled datasets for CNN classification. Label encoding was applied to convert string labels into integers.

### 2.7 Convolutional Neural Network classification of partial MPXV genomes

One-hot encoded genome from windows of *Length* = 1000 base pairs were classified using a hierarchical two-step CNN ensemble to improve classification amidst strong class imbalance. CNNs were implemented in the TensorFlow/Keras (version 2.19.0) library running on a high-performance computing cluster provided by the Digital Research Alliance of Canada (ACENET, Eastern Canada) on a computational node consisting of 4 CPUs per task, 128Gb of RAM memory, and an NVIDIA P100 Pascal (12G HBM2 memory) GPU. In the first stage, a CNN was trained to discriminate between the majority class (clade II) and the pooled minority classes (subclade Ia and subclade Ib). Samples predicted as belonging to the minority group proceeded to the second stage, in which an additional CNN was trained to differentiate subclade Ia from subclade Ib. Each model was implemented as a sequential architecture comprising multiple one-dimensional convolutional layers with progressively increasing filter counts (32-128) and kernel sizes of 5-7 followed by batch normalization a LeakyReLU activation functions. Max-pooling layers (size = 2) were interleaved to reduce special dimensionality, and dropout layers (rate = 0.3 – 0.5) were used to mitigate overfitting, The output layer of each model consisted of a dense softmax classifier with two output units corresponding to their binary task. Both models were trained using the Adam optimizer (learning rate = 1.10^-4^) and categorical cross-entropy loss. Early stopping was applied on the test loss with a patience of three epochs. The models were allowed to run for 50 training epochs each. The best models were saved as HDF5 files for downstream application.

### 2.8 Web development

We developed a user-friendly web server for interested users to execute the pretrained models for MPXV clade assignment using their own data. The web server was developed using Django (version 5.0.3). The Django web server is structured using a Model-View-Controller design pattern. The backend was developed using Python (version 3.12). No database is required for the application to run, consequently, no user information or data is ever stored. The frontend interface was built using HTML 5, CSS, JavaScript,and Bootstrap. The web server was deployed on a remote server containing Ubuntu 22.04 located in Toronto, Canada. Gunicorn serves as the WSGI server and Ajax as a reverse proxy directing the HTTPS requests to a domain name obtained within Dalhousie University.

### 2.9 Benchmarking CladePredictor-MPXV with other clade assignment tools

We compared the runtime performance of CladePredictor-MPXV and Nextclade for clade assignment on individual sequences from our validation dataset. Each sequence was processed individually by both tools. Both applications were executed using their respective Docker images on an Apple MacBook Pro (2022 M1 Pro, 32 GB RAM, macOS Venture 13.0). The Docker commands for each tool were wrapped in bash scripts that invoked GNU-time (version 1.9) to measure execution time and memory usage per genome. For Nextclade, we used the ‘mpox’ database as the reference dataset. Since both tools ran within Docker containers, which introduces system overhead, we measured baseline memory consumption by running an empty container with the same mounted volumes but no workload. This baseline was subtracted from the memory usage reported during actual runs to approximate the true memory consumption of each tool. Genome Detective is another tool that is capable of assigning clades to MPXV genomes. However, the tool is not free or open source. Hence, we did not consider it in our benchmarking.

## RESULTS

### 3.1 Trinucleotide frequency 2D visualization of MPXV genomes

To visualize genomic trinucleotide frequencies, we performed UMAP-based dimensionality reduction from 64 to 2-dimensions for genomes from MPXV subclades Ia, Ib, and IIb. By iterating over UMAP parameters, the best Silhouette score (0.44) was achieved with the UMAP parameters n_neighbors = 390 (10% of the data), min_dist = 0.7, metric = manhattan. The UMAP-based dimensionality reduction technique resulted in a well-defined subclade IIb cluster and subclade Ia and Ib sequences forming another cluster with a little overlap and many observations of subclade Ib being separated from subclade Ia (**Figure 1A**). Following the interpretation of **Figure 1A**, where trinucleotide frequencies are suitable for creating a major subclade IIb cluster, and a second cluster containing subclade Ia and Ib sequences with some overlap, we conducted a second step of UMAP isolating subclades Ia and Ib sequences (**Figure 1B**). With the parameters n_neighbors = 120, min_dist = 0.3, and metric = cosine, a Silhouette score of 0.23 was achieved was the UMAP dimension reduction rendered more distinct clusters of subclades Ia and Ib.

**Figure 1.**
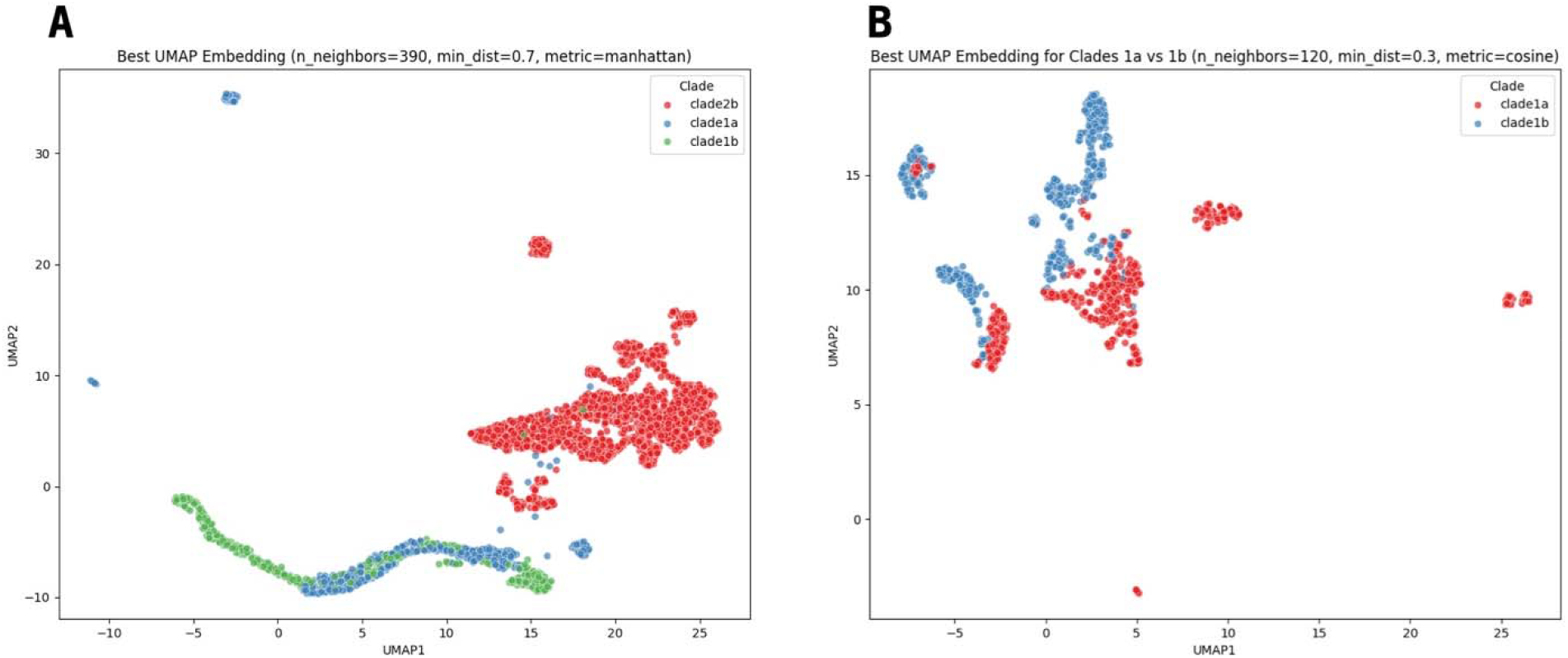
2D visualization of trinucleotide frequency of MPXV genomes. UMAP visualization of MPXV sequences from different clades (**Figure 1A**) and subclades (**Figure 1B**). A data matrix was generated containing the kmer (k=3) frequency of ATCG bases in the genomes of MPXV subcaldes Ia, Ib, and IIb. The shown UMAP plots represent the best Silhouette score in an iterative process over n_neighbors (1%, 5%, and 10% the number of observations), min_dist (0.1, 0.3, 0.5, 0.7, 1), and metric (euclidean, cosine, manhattan).

### 3.2 Complete genome classification with XGBoost

We used XGBoost to classify complete MPXV genomes based on trinucleotide frequencies. The classification task was split into two binary classifiers as indicated by the UMAP dimension reduction (**Figure 1**). First, we evaluated the complete genome classification model distinguishing between clade I and clade I on both the held-out test set, and the independent validation set (**Figure 2A**). On the test set (n = 564 sequences), the model demonstrated excellent classification performance, achieving an overall accuracy of 100%. Precision, recall, and F1-scores for both classes were near perfect, with clade I reaching a recall of 1 and clade II achieving perfect precision, recall, and F1-score. In contrast, performance on the validation set (n = 665 sequences), which included sequences from smaller phylogenetic clusters not seen during training, showed a decrease in accuracy to 86.7%. The model maintained high precision for clade I (1) but with reduced recall (0.68) indicating some false negatives within this class. Clade II sequences were predicted with balanced performance, achieving 0.81 precision and perfect recall, resulting in a F1-score of 0.90. These metrics suggest that while the classifier generalizes well to novel data, it tends to miss some clade I sequences. Next, we evaluated the complete genome classification model distinguishing between the subclades Ia and Ib (**Figure 2B**). On the test set (n=176 sequences), the model achieved an accuracy of 98.2%. The precision and recall were well-balanced across the classes: subclade Ia exhibited a high recall of 1 and precision of 0.96, indicating that most subclade Ia sequences were correctly identified. Subclade Ib had slightly higher precision (1) but a lower recall (0.97), showing the model was very accurate when predicting subclade Ib but missed a small portion of true subclade Ib sequences. The F1-scores were similarly high for both classes, reflecting a good balance between precision and recall. On the validation set (n=278 sequences), which included independent sequences not seen during training, the classifier maintained its robustness with an accuracy of 98.2%. Precision and recall remained high and consistent with the test set results: subclade Ia with 0.98 precision and 0.99 recall while subclade Ib with 0.98 precision and 0.97 recall. The model demonstrated strong generalization capacity, with minimal loss in sensitivity or specificity on new data.

**Figure 2.**
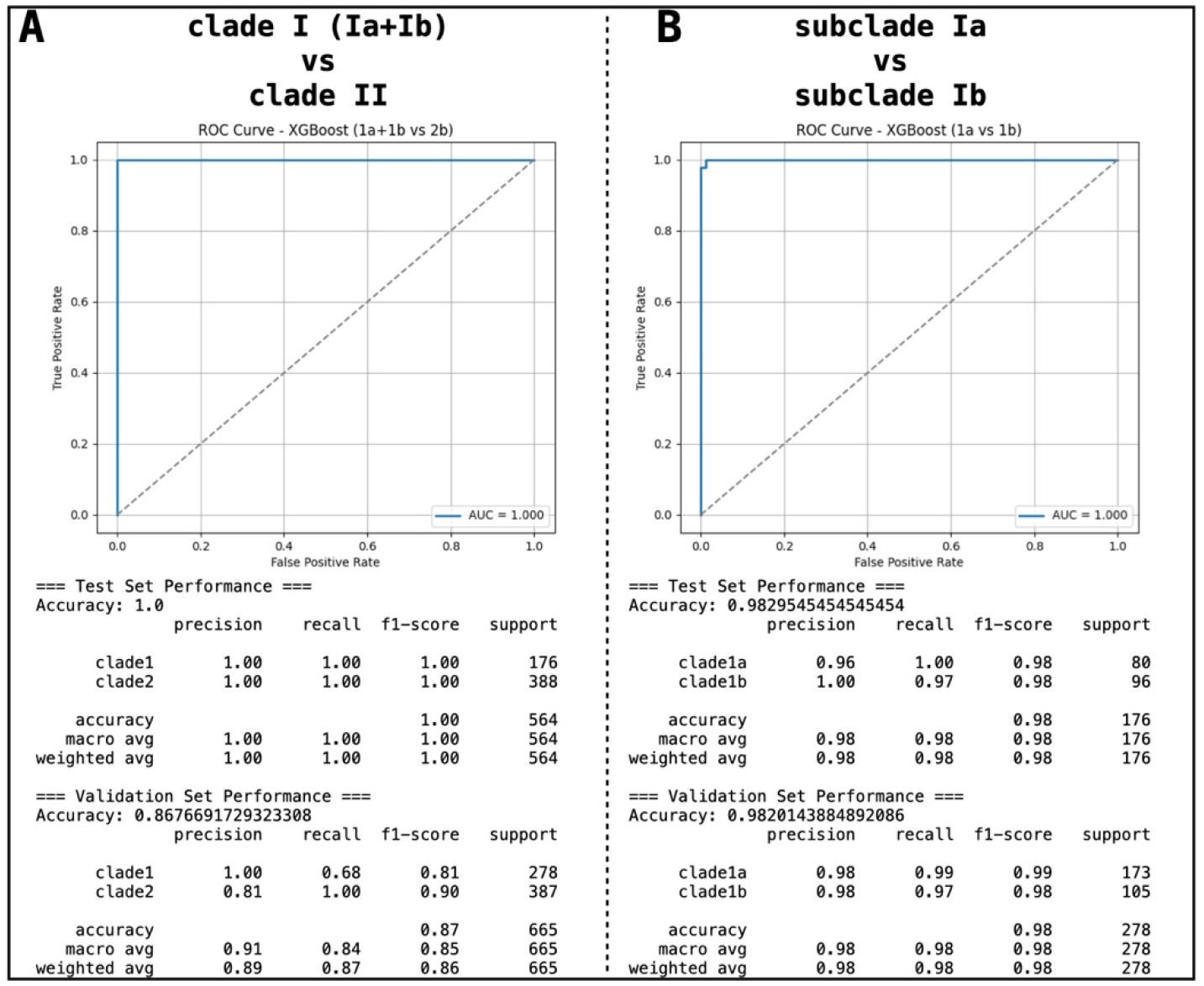
XGBoost classifiers of MPXV Clade (I vs II) and subclades (Ia vs Ib) complete genomes.

In **Figure 2A** we show the ROC curve for the XGBoost classifier distinguishing clade I from clade II sequences. The model achieved a perfect area under the curve (AUC = 1), indicating excellent discrimination between the two classes with no classification errors on the test set. Below, we show the model overall model performance both on the test and validation sets. In **Figure 2B**, we show the ROC curve for the XGBoost classifier distinguishing between subclades Ia and Ib. The model achieved almost perfect area under the curve score (AUC = 1), indicating excellent discrimination between the two classes. Below, we show the overall performance of the model both on the test and validation sets.

We also evaluated the performance of XGBoost models in classifying partial MPXV genomes. We derived partial genomes from the complete genomes in windows *w* starting with a size = 196000 and decreasing by 1000 and generated multiple MPXV partial genomes (Methods 2.5). Each generated partial genome was submitted to the ensemble of XGBoost classifiers and the performance was evaluated on each length. Overall, as the genomes length decrease, the performance of our model also decreases, dropping below 0.9 accuracy for input genomes with a length less than 188,000 base pairs. This poor performance motivated the development of an alternative approach for the classification of partial genomes, i.e., genomes with a length less than 188,000 base pairs. The individual performance of the ensemble of XGBoost classifiers for each genome length is available in **Supplementary Material S2**.

### 3.3 Partial genome classification with CNNs

To classify partial MPXV genomes, we generated overlapping 1000-nucleotides windows with a step size of 500-nucleotides step all genomes in the train, test, and validation data (methods 2.6), which consist of complete genomes. Using convolutional approaches to capture local feature dependencies, we trained an ensemble of two CNNs with binary classification tasks: clade I vs. clade II and subclade Ia versus subclade Ib. The CNN model for clade I versus clade II classification trained for ten epochs, achieving its optimal performance at epoch six, where it attained the lowest test loss of 0.0216 and a test accuracy of 99.72%. Similarly, the CNN model for subclade Ia versus subclade Ib classification underwent eight training epochs, with the best generalization performance observed at epoch five with a minimum test loss of 0.3121 and a test accuracy of 85.42% (**Figure 3A**). In the first stage, a binary CNN distinguished the majority class (clade II) from the minority class (subclade Ia and Ib, combined). When evaluated on the independent validation set, the full ensemble achieved an overall accuracy of 95% with a macro-averaged F1-score of 0.88 demonstrating balanced prediction across all classes. Subclade Ia achieved a precision of 0.77 and a recall of 0.88, while subclade Ib reached precision of 0.88 and recall of 0.79. The majority class (subclade IIb) was classified with perfect precision and recall, demonstrating the effectiveness of the binary gating stage (**Figure 3B**). Ultimately, we constructed partial MPXV genomes by combining random 1000-nucleotide windows from the genomes in the validation set (>1 window and < than the total number of windows per genome). Each window had a CNN prediction associated with it (subclade Ia, Ib, or IIb). For a set of *n* windows, we tracked the majority of the assigned predictions and determined it as the final prediction of the partial genome. If there is a tie between any of the predicted classes, the partial genome is listed as undetermined. Our validation dataset is composed of 623 genomes out of which 590 could generate at least one 1000-nucleotide window. Out of the 97 subclade Ia partial genomes, 88 were correctly predicted (four predicted as subclade Ib, three as undetermined, and two as IIb). Out of the 105 subclade Ib partial genomes, 96 were correctly predicted (eight as subclade Ia, one as undetermined). Out of the 387 subclade IIb genomes, all the 387 were correctly predicted (**Figure 3C**). Even in instances where the partial genome was composed of less than 10 windows (27 occasions), the correct subclade was predicted in 18 occasions.

**Figure 3.**
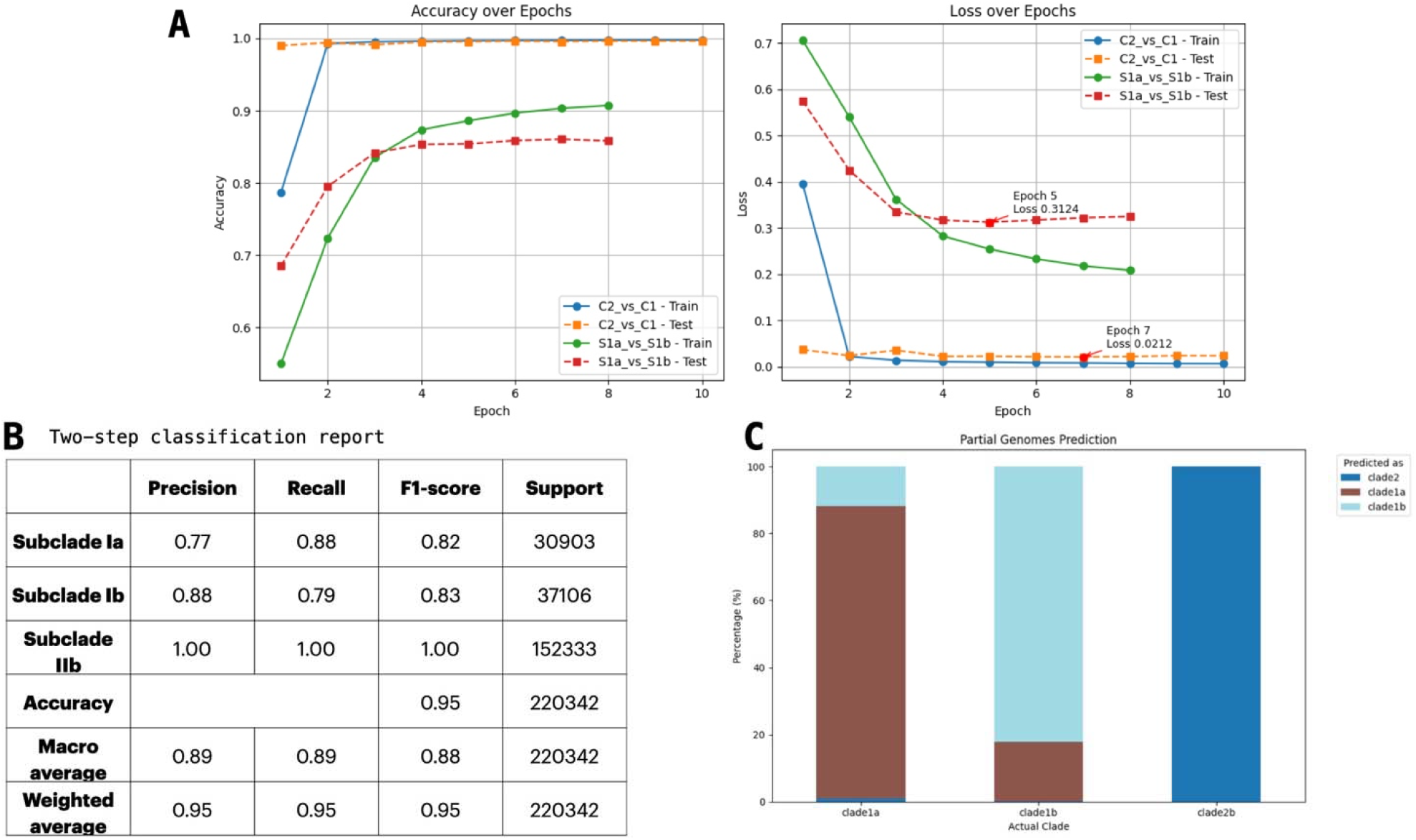
Convolutional Neural Network classification of MPXV partial genomes. We show in **Figure 3A**, the train/test accuracy and loss (cross entropy function) over the training epochs of the CNN ensemble of classifiers of MPXV clades (I vs II) and subclades (Ia vs Ib). Initially, the total number of epochs was set to 50. An early stopping was added where if the validation loss does not decrease after three consecutive epochs, the training stops. After stopping, the model restores the weights from the epoch with the lowest validation loss and is saved for downstream use. In **Figure 3B**, we show the combined classification report for the two binary classification tasks. We display class-specific precision, recall, and F1 score values. We also checked the accuracy of the model in classifying each clade in a macro average without compensating for class imbalance and weighted average. In **Figure 3C**, we show the validation step of the classification where the complete genomes from the validation set were split into windows of 1000 nucleotides in a 500-nucleotide step. For each genome, a random *n* number of windows were selected where *n*> 1 and *n* < *total_windows*. Each window was submitted to the CNN partial genome classifier. The total number of windows per subclade is show in the y-axis as percentages. The bars are colored according to the number of windows predicted as subclade Ia, Ib, or IIb.

### 3.4 CladePredictor-MPXV: An AI-ensemble classifier of complete and partial MPXV genomes

CladePredictor-MPXV implements an ensemble of classifiers that can accurately predict clade and subclade labels for complete (using global features) and partial genomes (using local features) of MPXV genomes. We represent the workflow of CladePredictor–MPXV in **Figure 4** in a schematic flowchart. Each query input MPXV genome is evaluate for length, if it is >= 188,000 base pairs, the trinucleotide frequency is calculated, and the genome is classified using the XGBoost clade classifier (output either clade I or clade II). If the XGBoost clade classifier classified the input genome as Clade I, CladePredictor-MPXV passes the trinucleotide frequency matrix into a second XGBoost classifier for subclade prediction into subclade Ia or subclade Ib. Alternatively, for partial input genomes that are less than 188,000 base pairs in length, CladePredictor-MPXV slices the input genome into *n* windows containing 1000 nucleotides each in a 500-nucleotide step size. The data matrix then sent to the hierarchical CNN partial genome classifier, where each window will first be predicted as clade I or clade II and if predicted as clade I, further predicted as subclade Ia or Ib. In both classifications, the majority of windows predicted to a subclade is the output. CladePredictor-MPXV is freely available at https://clade-predictor.microbiologyandimmunology.dal.ca. We also provide a containerized application of CladePredictor-MPXV for local execution at https://hub.docker.com/r/sganzg/clade_predictor

**Figure 4.**
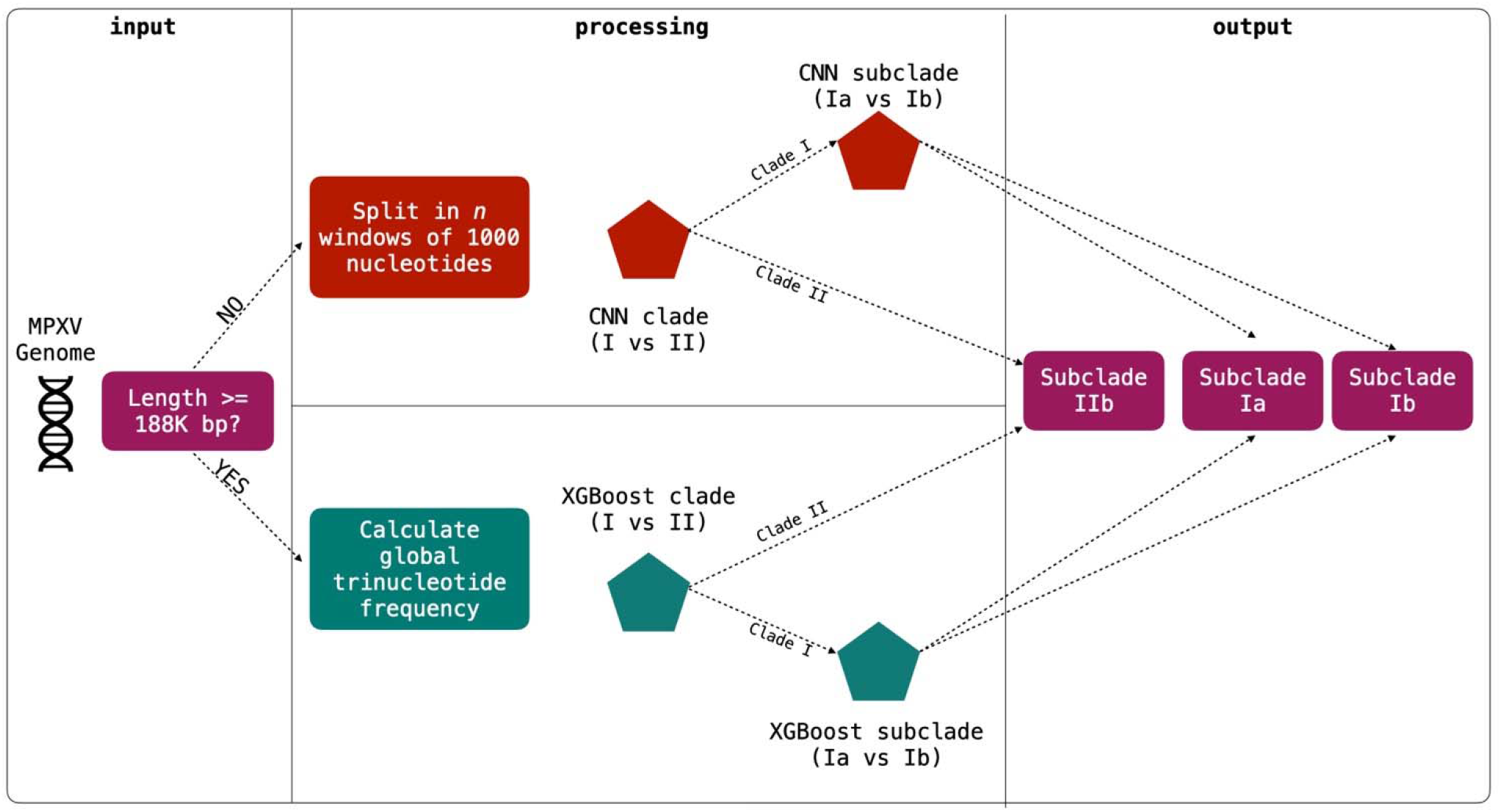
CladePredictor-MPXV classification workflow. CladePredictor-MPXV classification flowchart depicting inputs, processing varying from different input lengths (complete and partial genomes), and output into subclade Ia, Ib, or IIb.

### 3.5 Benchmarking CladePredictor - MPXV and NextClade

We ran individual MPXV genomes from our validation in CladePredictor-MPXV and Nextclade to assess their time and memory consumption. The clade assignment task of CladePredictor-MPXV took on average 2.7±0.11 (median=2.68, IQR=2.63-2.75) seconds while Nextclade took on average 7.14±1.29 (median=6.78, IQR=6.49-8.31) seconds (**Figure 5A**). The overhead RAM memory usage of CladePredictor-MPXV was on average 5.8±28.46 (median=0, IQR=0-0) KB while Nextclade consumed on average 114.62±156.48 (median=32, IQR=0-192) KB (**Figure 5B**). In conclusion the average time and overhead RAM memory consumption of CladePredictor-MPXV is significantly lower than Nextclade in individually processing complete MPXV genomes. It is worth noting that the classification process of CladePredictor-MPXV had no measurable increase over baseline memory usage in 631 out of 665 genomes.

**Figure 5.**
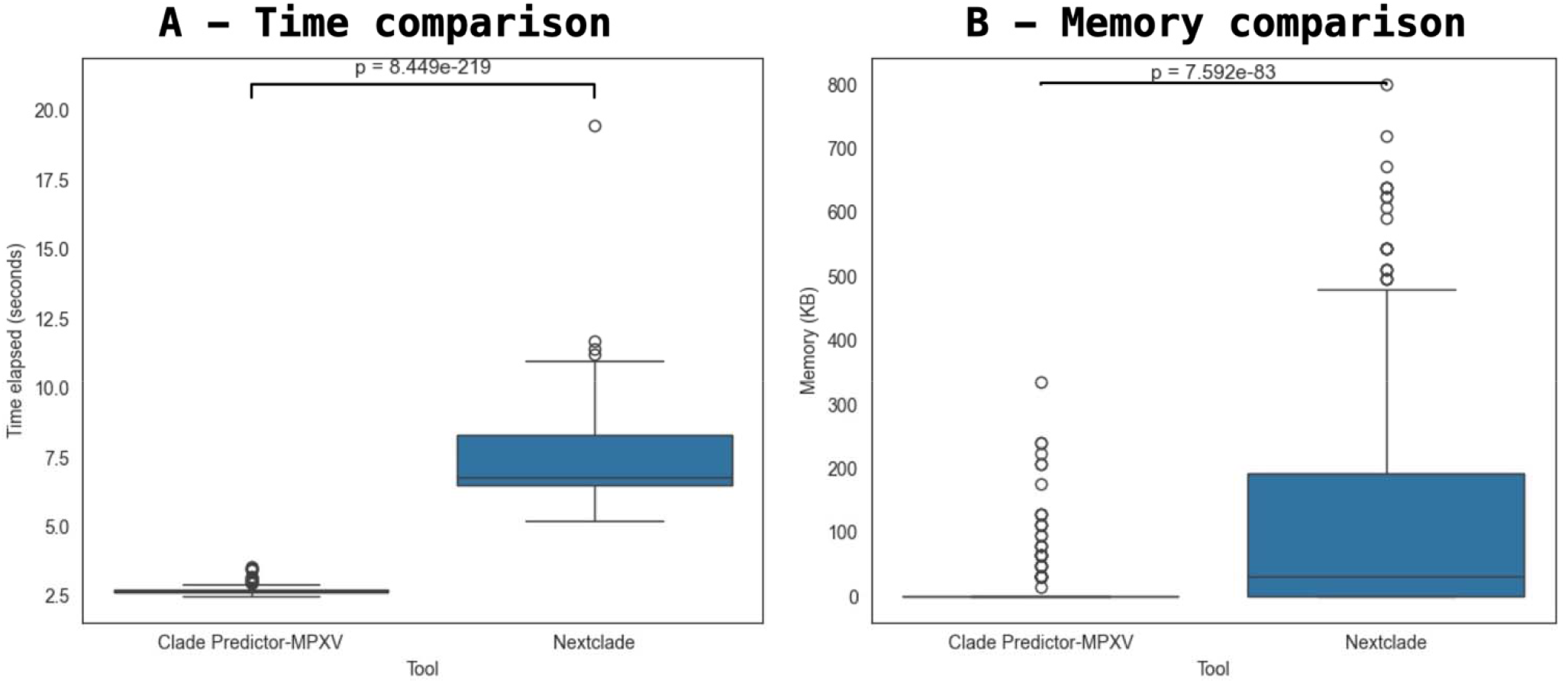
Time and overhead memory consumption for CladePredictor-MPXV and Nextclade to process individual MPXV genomes.

In **Figure 5A**, the run time (measured as time elapsed, seconds) of CladePredictor-MPXV and Nextclade was compared in individually processing 665 MPXV genomes. The variable ‘time elapsed was found as non-parametric (Shapiro-Wilk *p* = 2.37e-36). We compared the average time consumed of both classifiers using the Mann-Whitney U test. The time consumption of CladePredictor-MPXV was significantly lower than of Nextclade (*p* = 8.44e-219). In **Figure 5B**, the RAM memory overhead (measured as the difference between the application running processing data and a baseline with no data) of CladePredictor-MPXV and Nextclade was compared. The variable ‘max resident set’ was found as non-parametric (Shapiro-Wilk *p* = 1.44e-49). The memory consumption of CladePredictor-MPXV was significantly lower than of Nextclade (*p* = 7.59e-83).

## DISCUSSION

Current MPXV classification methodologies often rely on whole-genome alignments, which can be computationally intensive. Alignment-free genome classification methods have been listed as efficient alternatives compared to traditional alignment-based methods [16]. As the volume of input data increases, alignment-based methods approaches might become impractical due to high memory and processing demands, limiting their scalability for large datasets. One widely used tool for the classification of MPXV genomes is Nextclade [8], which uses an implementation of the Smith-Waterman alignment [17] and can correctly classify MPXV sequences. On the other hand, Kraken2 [10], which is a taxonomic classifier of metagenomics data, can also classify genomes and utilizes kmer frequencies as input. Despite k-mer approaches being computationally appealing ways to treat genomic information, information about the order and relationship of genomic segments is lost, which might be biologically relevant. To overcome some of the inherent challenges associated with sequence-based classification, we developed a hybrid tool that integrates distinct ML models for the classification of complete and partial MPXV genomes into subclades Ia, Ib, and IIb. Our approach uses the complementary strengths of a k-mer-based feature extraction and CNNs to capture both global and local sequence features.

The complete genome classifier of CladePredictor-MPXV was not able to capture MPXV Clade II genomes with less than 188,000 nucleotides. This further led us to create a classifier based on local features to classify partial genomes. Given that XGBoost and gradient boosting decision trees classifiers will capture features in a global space and use them for classification, it is expected that its efficiency in handling structured, high-dimensional datasets makes it a strong classifier for tasks involving comprehensive feature vectors derived from entire genomes. Evidence of XGBoost’s robustness with global scale data is found in the study from Xia et al, (2024) [18] where the authors used XGBoost to process datasets with extensive feature spaces. The model demonstrated better classification performance in scenarios that required the analysis of the interaction of global features. The classification framework of CladePredictor-MPXV benefits from decomposing multiclass classification into an ensemble of binary classifiers of observations. The current iteration of our tool is only able to distinguish between the subclades Ia, Ib, and IIb of MPXV as they contain most of the deposited genomes. Although our initial design did not encompass other organisms and less represented subclades of MPXV, our classification framework can be adapted and translated to accommodate the subtyping of other organisms.

In conclusion, we believe the scientific community has a lot to benefit from an alignment-free clade classifier of MPXV genomes, given the global health impact mpox has caused over the past few years. The scalability of our work advocates for the development of additional artificial intelligence models that can capture the distinctive genetic profile to discern other viruses and microorganisms in general. Our hierarchical, dual-mode classification framework represents an efficient tool for the rapid assignment of clades to both complete and incomplete genome assemblies, accommodating the variable quality and fragmentation frequently encountered in metagenomic and outbreak surveillance data. CladePredictor-MPXV is freely available at https://clade-predictor.microbiologyandimmunology.dal.ca.

## Data Availability

The findings of this study are associated with MPXV genome sequences deposited on GISAID via EPI_SET_250815fu, which are accessible at https://doi.org/10.55876/gis8.250815fu. We appreciate and recognize the efforts of the originating laboratories in providing sequences. CladePredictor-MPXV is open-source and it's source code is publicly available at https://github.com/gustavsganzerla/mpxv_clade_prediction

## Acknowledgements

We thank Dr. Nikki Kelvin for critical proofreading and editing of this manuscript. We also thank the Digital Research Alliance of Canada and ACENET, their provider of high-performance computing in the Atlantic Canada, for providing the computational infrastructure necessary to train the models. The findings of this study are associated with MPXV genome sequences deposited on GISAID via EPI_SET_250815fu, which are accessible at https://doi.org/10.55876/gis8.250815fu. We appreciate and recognize the efforts of the originating laboratories in providing sequences.

